# Distribution of White Matter Hyperintensities across Arterial Territories in Neurodegenerative Diseases

**DOI:** 10.1101/2024.09.29.24314328

**Authors:** Ikrame Housni, Flavie E. Detcheverry, Manpreet Singh, Mahsa Dadar, Chloe Anastassiadis, Ali Filali-Mouhim, Mario Masellis, Zahinoor Ismail, Eric E. Smith, Simon Duchesne, Maria Carmela Tartaglia, Natalie A. Phillips, Sridar Narayanan, AmanPreet Badhwar

## Abstract

MRI-detected white matter hyperintensities (WMH) are often recognized as markers of cerebrovascular abnormalities and an index of vascular brain injury. The literature establishes a strong link between WMH burden and cognitive decline, and suggests that the anatomical distribution of WMH mediates cognitive dysfunction. Pathological remodeling of major cerebral arteries (anterior, ACA; middle, MCA; posterior, PCA) may increase WMH burden in an arterial territory (AT)-specific manner. However, this has not been systematically studied across neurodegenerative diseases (NDDs). To address this gap, we aimed to assess WMH distribution (i) across ATs per clinical category, (ii) across clinical categories per AT, and (iii) between men and women. We also investigated the association between AT-specific WMH burden and cognition.

Using two cohorts – Canadian CCNA-COMPASS-ND (N=927) and US-based NIFD (N=194) – we examined WMH distribution across ten clinical categories: cognitively unimpaired (CU), subjective cognitive decline (SCD), mild cognitive impairment (MCI), Alzheimer disease (AD), MCI and AD with high vascular injury (+V), Lewy body dementia, frontotemporal dementia, Parkinson’s disease (PD), and PD with cognitive impairment or dementia. WMH masks were segmented from FLAIR MRI and mapped onto an arterial atlas. Cognitive performance was assessed using four psychometric tests evaluating reaction time and overall cognition, namely Simple Reaction Time (SRT), Choice Reaction Time (CRT), Digit Symbol Substitution Test (DSST), and Montreal Cognitive Assessment (MoCA). Statistical analyses involved linear regression models, controlling for demographic factors, with a 5% False Discovery Rate for multiple comparisons.

Our transdiagnostic analysis revealed unique AT-specific WMH burden patterns. Comparisons between ACA and PCA territories revealed distinct burden patterns in clinical categories with similar whole-brain WMH burden, while the MCA territory consistently exhibited the highest burden across all categories, despite accounting for AT size. Hemispheric asymmetries were noted in seven diagnostic categories, with most showing higher WMH burden in the left MCA territory. Our results further revealed distinct AT-specific WMH patterns in diagnostic groups that are more vascular than neurodegenerative (i.e., MCI+V, AD+V). Categories often misdiagnosed in clinical practice, such as FTD and AD, displayed contrasting WMH signatures across ATs. SCD showed distinct AT-specific WMH patterns compared to CU and NDD participants. Additionally, sex-specific differences emerged in five NDDs, with varying AT effects. Importantly, AT-specific WMH burden was associated with slower processing speed in MCI (PCA) and AD (ACA, MCA).

This study highlights the importance of evaluating WMH distribution through a vascular-based brain parcellation. We identified ATs with increased vulnerability to WMH accumulation across NDDs, revealing distinct WMH signatures for multiple clinical categories. In the AD continuum, these signatures correlated with cognitive impairment, underscoring the potential for vascular considerations in imaging criteria to improve diagnostic precision.

## 1. INTRODUCTION

The world population is aging, and the rise of age-related neurodegenerative diseases (NDDs) poses a major health challenge [1–3], with over 55 million people affected [4,5]. Of the NDDs, Alzheimer’s disease (AD) is the most prevalent [6,7]. Mounting evidence indicates a pivotal role of cerebrovascular pathology in the initiation, progression, and/or exacerbation of NDDs [8–11]. Cerebrovascular pathology features routinely detected by non-invasive *in vivo* magnetic resonance imaging (MRI) are white matter hyperintensities (WMH), covert brain infarcts, microbleeds, lacunes, and enlarged perivascular spaces, with WMH being the most common [12,13]. A recent meta-analysis investigating these four features found only WMH burden to be significantly associated with increased risk of dementia (including AD) in the general population [12].

WMH appear as areas of elevated signal in white matter on T2-weighted, proton density-weighted, or fluid-attenuated inversion recovery MRI [14]. Increased water content in brain white matter due to axonal damage (demyelination, loss), and to some extent edema, are thought to contribute to the appearance of WMH [15,16]. Putative vascular pathogenic mechanisms of WMH include endothelial and associated blood-brain-barrier dysfunction, hypoperfusion due to altered cerebrovascular autoregulation and reactivity, and cerebral amyloid angiopathy [16–25].

Emerging literature points to NDD-specific spatial distribution of WMH in the brain [26]. Examples include a significantly higher WMH burden in the (i) deep [27] and right-posterior [28] brain regions in vascular dementia compared to AD, and (ii) periventricular and subcortical regions in AD compared to Parkinson’s disease (PD) [29]. Studies further suggest that these NDD-specific spatial distributions of WMH can impact cognitive function [11,26,29,30]. To date, the main analytical approaches utilized to map NDD-specific WMH topography consist of: region of interest (ROI)-based approaches, with most studies investigating WMH distribution in cortical lobes (e.g., frontal versus parietal); ventricular-based approaches, with studies assessing WMH distribution based on distance from ventricular surface (e.g., periventricular versus deep); and voxel-based approaches, with studies assessing presence of WMH in each white matter voxel [26]. Lacking, however, is an appreciation of the relationship of WMH location with respect to a cerebrovascular architecture (e.g., arterial-territory, -density) informed mapping.

Indeed, accumulating observations collectively point to a crucial need for cerebral arterial-territory (AT) informed mapping of NDD-specific distributions of WMH. Most of the brain, namely the supratentorial brain, is largely served by blood vessels arising from three major cerebral arteries (anterior, ACA; middle, MCA; posterior, PCA), properties of which can give rise to region-specific effects. For instance, lumen diameter and wall thickness of these arteries have been shown to correlate positively with lumen diameter of downstream ipsilateral arterioles [31]. Physiological (e.g., aging) or pathological remodeling of these arteries, therefore, has the potential to stimulate AT-specific remodeling of distal arterioles [31] and increase WMH burden [32]. In fact, cerebral arteries comprising the anterior (including ACA and MCA [33,34]) and posterior (including PCA [33,34]) circulation [35] differ in the distribution of age-specific histopathological features (e.g., elastin loss, increased collagen and/or amyloid-beta (Aβ) deposition), with dissimilarity in arterial properties of the two circulations (e.g., sympathetic innervation) as a contributing factor [36,37]. Moreover, emerging studies demonstrate that the spatial patterning of age-specific cerebrovascular pathology may modulate NDD-specific risk-factors and/or changes (e.g., *APOE-E4* allele, increased Aβ deposition) [38–42]. For example, age-related increase in collagen content in posterior relative to anterior circulation promotes Aβ accumulation, which may contribute to increased cerebral amyloid angiopathy in the posterior circulation [35,40] and the posterior predominance of WMH in AD [43].

To date, while AT-informed mapping of WMH distribution is lacking for NDDs, two non-NDD studies highlight the utility of conducting such investigations. The first study found that in cognitively unimpaired (CU) older adults (60-64 years of age), subregions of the ACA and MCA territories, supplied by the medial and lateral lenticulostriate arteries, respectively, exhibited the highest prevalence of WMH [44]. The second study investigated stroke, itself a risk factor for NDD [45] found that ACA WMH burden was most associated with stroke risk-factors over MCA and PCA [46].

In the present study, we conducted **AT-informed mapping of NDD-specific WMH distributions**, and hypothesized the presence of NDD-specific signatures. Our three main aims were to assess WMH volume (i) across ATs within each clinical category (Aim 1), (ii) across clinical categories per AT (Aim 2), and (iii) between men and women per clinical category and AT, given that sex-specific differences in WMH volume have been reported [47] (Aim 3). Given that both increased WMH burden [48] and its anatomical distribution [11] mediate cognitive dysfunction, a follow-up aim (Aim 4) was to investigate the association between AT-specific WMH burden and cognition.

## 2. METHODS

### 2.1. Participants

#### CCNA COMPASS-ND cohort

We used participant data (demographic, cognition, cross-sectional MRI) from the Comprehensive Assessment of Neurodegeneration and Dementia (COMPASS-ND) cohort of the Canadian Consortium on Neurodegeneration in Aging (CCNA), a pan-Canadian initiative to catalyze and promote dementia-related research [49]. The 927 participants included were distributed across ten clinical categories, namely, CU (N=107), as well as subjective cognitive impairment or decline (SCD; N=133), mild cognitive impairment due to AD (MCI; N=240; i.e., the prodrome stage of AD), MCI with a high vascular brain injury (MCI+V; N=133), AD (N=93), AD with a high vascular brain injury (AD+V; N=46), Lewy body dementia (LBD; N=26), frontotemporal dementia (FTD; N=23), PD (N=77), and PD with cognitive impairment or dementia (PDD; N=49). Per participant, clinical diagnosis was determined by physicians based on standard diagnostic criteria following evaluation of information from recruitment assessment, screening visit, longitudinal clinical visits, and MRI [49]. Ethical agreements and written informed consents were obtained from all participants [49,50]. Information on availability and access to CCNA COMPASS-ND data can be requested at https://ccna-ccnv.ca/contact/.

#### NIFD cohort

Due to the low CCNA COMPASS-ND FTD sample size, we also included FTD participants from the Frontotemporal Lobar Degeneration Neuroimaging Initiative (FTLDNI also known as NIFD) to validate the replicability of our findings. The NIFD is a multi-site study funded through the National Institute of Aging that aims to identify neuroimaging biomarkers of FTD. In addition to our CCNA COMPASS-ND analyses, we conducted separate analyses on the sample collected at the primary collection site, which consisted of 194 participants distributed across two clinical categories, namely, cognitively unimpaired (CU_NIFD; N=96) and frontotemporal dementia (FTD_NIFD; N=98). Access to the dataset can be requested from Laboratory of Neuro Imaging (LONI) (https://ida.loni.usc.edu/login.jsp) and further information can be found at https://4rtni-ftldni.ini.usc.edu/.

#### Naming convention used for the CCNA COMPASS-ND and NIFD cohorts

Our data included two FTD and CU groups from the CCNA COMPASS-ND and NIFD datasets. We use the terms "FTD" and "CU" specifically for groups from the CCNA COMPASS-ND cohort. For groups from the NIFD cohort, we use the terms "FTD_NIFD" and "CU_NIFD." This naming convention ensures clear differentiation between the cohorts throughout our analysis.

### 2.2. Magnetic Resonance Imaging Data

#### CCNA COMPASS-ND cohort

All CCNA COMPASS-ND participants were scanned following the Canadian Dementia Imaging Protocol (CDIP; [51]), a harmonized MRI protocol designed to reduce inter-scanner variability in multi-centre studies. MRI was performed using three 3 tesla (T) scanner models, namely GE, Philips, and Siemens. MRI sequences used to detect WMH included 3D isotropic T1-weighted and 2D T2-weighted fluid attenuated inversion recovery (slice thickness 3mm, FLAIR: TR 9000 ms, TI 2500) acquisitions.

#### NIFD cohort

3D T2-weighted FLAIR scans (slice thickness 1mm, TR 6000 ms, TE 389 ms, inversion time 2100 ms) and MPRAGE T1-weighted scans (slice thickness 1mm, TR 2300 ms, TE 3ms, TI 900 ms) were obtained on a 3T Siemens TrioTrim scanner for all participants, following a harmonized protocol based on Alzheimer’s Disease Neuroimaging Initiative (ADNI) recommendations [52].

##### 2.2.1 MRI-preprocessing and WMH-segmentation

###### CCNA COMPASS-ND cohort

MRI scans were downloaded from the CCNA COMPASS-ND LORIS database [53]. Image preprocessing, including (i) image denoising [54], (ii) correction for intensity non-uniformity [55], and (iii) normalization of image intensities (set range: 0-100), was performed as previously described [56]. FLAIR images were co-registered to the T1-weighted images using a 6-parameter rigid-body registration and mutual information cost function [56]. Images were visually assessed for artifacts to ensure successful registration before being included in our analyses.

WMH were segmented in native FLAIR space using a publicly available automated segmentation method [57–59]. All segmentations were visually quality controlled to ensure their accuracy. T1-weighted images were linearly registered (9 parameters) to the MNI-ICBM152-2009c template space using a cross-correlation objective function [56]. To adjust for brain size variations and enable atlas-based regional analyses, MRI images and WMH maps were linearly transformed to MNI standard space. WMH burden was assessed per ROI using an AT atlas [46] in standard space (Fig. 1).

**Fig. 1.**
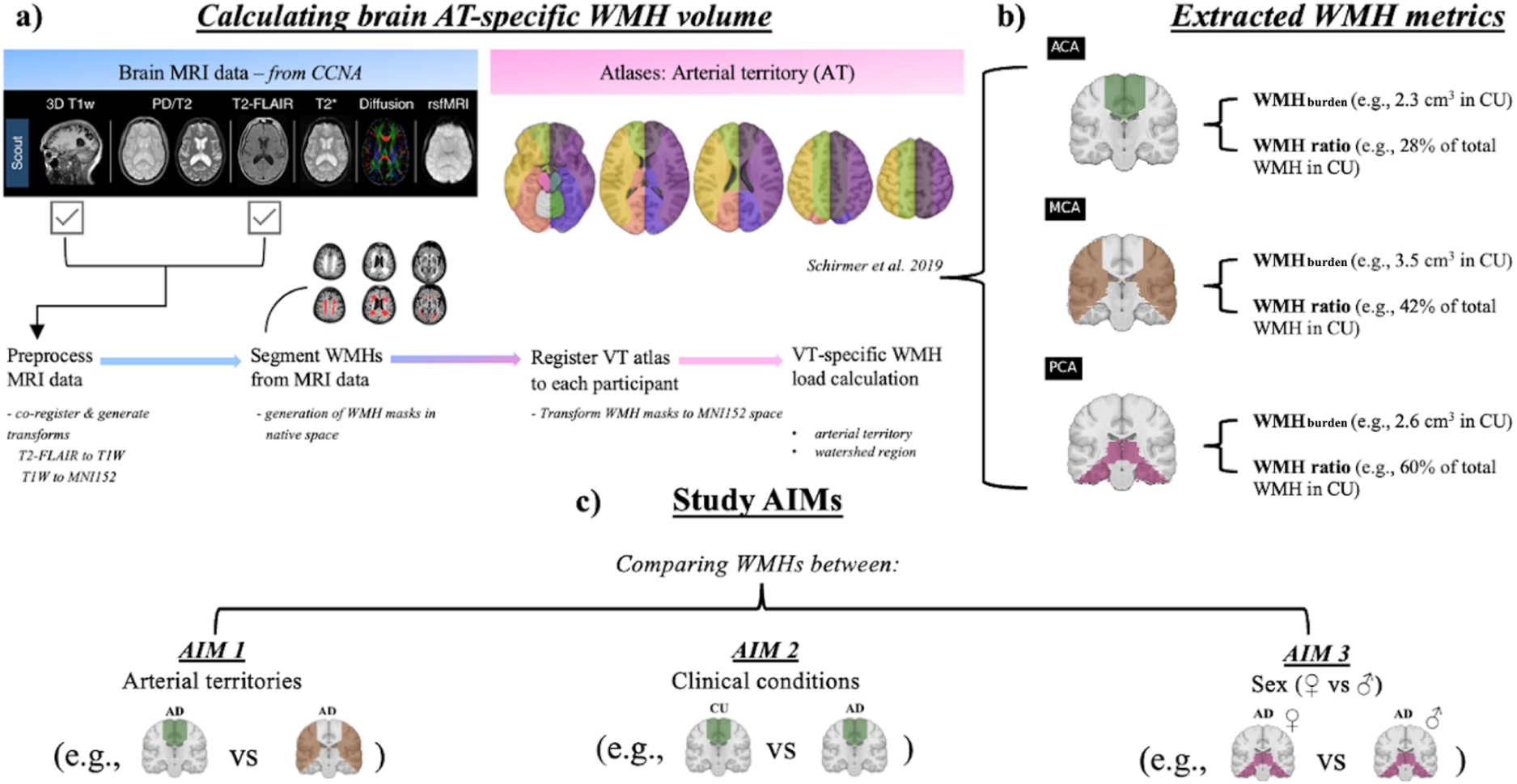
**a-b)** Workflow for calculating AT-specific <u>WMH metrics</u>: regional *WMH burden*, *WMH ratio* (regional burden/whole-brain burden) – examples provided. **c)** Aims of completed study; *Aim 1*: comparing WMH burden across ATs, per clinical category, *Aim 2*: comparing AT-specific WMH metrics across clinical categories, *Aim 3*: comparing WMH metrics across sex per clinical category and AT.

###### NIFD cohort

WMH were segmented using the open-access Lesion Prediction Algorithm (LPA) from the Lesion Segmentation Toolbox (LST) [60] available for download at https://www.applied-statistics.de/lst.html, and run on SPM12 (Matlab 2020). The resulting lesion probability maps were thresholded at 0.4 and manual corrections were performed by C.A. (blinded to diagnosis) to improve segmentation quality in ITK-SNAP 3.8.0. [61] (http://www.itksnap.org/pmwiki/pmwiki.php?n=Main.HomePage). WMH maps were then spatially normalized to the ICBM152 linear template using SPM12. The WMH burden was assessed per ROI using an AT atlas [46] in standard space.

##### 2.2.2 Investigated ROIs

We delineated ten ROIs from the supratentorial brain based on their vascular supply (Fig. 1a-b). Specifically, we concentrated our analysis on the major cerebral arteries, namely the ACA, MCA, and PCA. This macro-level approach allowed us to explore the three major networks of brain arteries. The analysis excluded the vertebro-basilar territory due to a significant decrease in data quality of FLAIR images in that region, rendering it insufficient for reliable use in our study. The included ROIs were grouped and statistically analyzed in three independent levels. **Level 1 (1 ROI):** *whole-brain –* supratentorial brain. **Level 2 (3 ROIs):** *bilateral ATs* – ACA territory (ACA_t_), MCA territory (MCA_t_), and PCA territory (PCA_t_). **Level 3 (6 ROIs):** *lateral ATs –* left ACA_t_ (LACA_t_), LMCA_t_, LPCA_t_, and right ACA_t_ (RACA_t_), RMCA_t_, RPCA_t_. Statistical analyses were performed and adjusted for multiple comparisons within each level.

##### 2.2.3 WMH metrics

Two metrics were used to assess the spatial distribution of WMHs, per ROI, namely WMH burden and WMH ratio. The regional WMH burden represents the volume of WMH within a given ROI. AT-specific WMH burden was corrected for AT-size to control for differences in region size when comparing inter-AT WMH burden [46]. The WMH ratio represents the proportion of regional WMH burden relative to the overall WMH burden, per ROI. This was mathematically assessed by dividing a ROI’s WMH burden by the whole-brain’s WMH burden.

Note that whenever we examined WMH in the whole-brain (i.e., Aims 2 and 3), we only evaluated the WMH burden, as the WMH ratio (which controls for the WMH burden of the entire brain) would invariably yield a value of 1 whenever the whole-brain is the ROI. Similarly, in Aim 1, we exclusively compared AT-specific WMH burden, as comparing AT-specific WMH ratios within the same individual would involve fractions with identical denominators. Consequently, conducting WMH ratio analyses in addition to WMH burden analyses in Aim 1 would have been redundant.

### 2.3. Statistical Analysis

#### 2.3.1 Analyses for AIMs 1-3: AT-specific WMH distributions

All statistical analyses were performed using R statistical software (v 4.1.3; R Core Team 2022) [62]. A series of generalized linear models (GLMs) were used with log-transformed region-size normalized WMH load as the dependent variable and clinical category, age, and sex as independent variables followed by appropriate pairwise comparisons of the WMH metrics. After fitting the models, we tested the validity of the linear model by assessing the normality of the residuals on the fit. To perform these comparisons, the *emmeans* function in R (emmeans; v 4.1.3; R Core Team 2022) used our GLMs as input to calculate the Estimated Marginal Means (EMMs) of our factor levels (predicted mean response of each level), while adjusting for the effects of the other factors in the model [63]. Subsequently, pairwise comparisons were conducted between the EMMs of each level to determine the significant differences between our factor levels. A 5% False Discovery Rate (FDR) was used to correct for multiple comparisons, adjusted *p*-values are reported.

As the CCNA COMPASS-ND and NIFD cohorts used different acquisition and imaging protocols, we performed analyses separately within each dataset. This approach is taken to avoid measurement and selection biases due to acquisition differences, ensuring the comparisons are reliable. Therefore, for group pairwise comparisons, all ten CCNA COMPASS-ND diagnoses will be compared against each other within the CCNA COMPASS-ND cohort, while in the NIFD cohort, only FTD_NIFD and CU_NIFD will be compared.

##### Inputted GLM models

*Aim 1. WMH burden per clinical category* ∼ *1 + **ROIs** + 1|Subject*

The model assessed differences in WMH burden between ROIs (variable of interest) for each clinical category, while ensuring subject pairing. Linear models and multiple comparison corrections were applied separately within CCNA COMPASS-ND and NIFD cohorts.

*Aim 2. WMH metric per ROI* ∼ *1 + **clinical category** + Sex + Age*

The model assessed differences in regional WMH burden or ratio between clinical categories (variable of interest) for each ROI, while correcting for age and sex. Linear models and multiple comparison corrections were applied separately within CCNA COMPASS-ND and NIFD cohorts.

*Aim 3. WMH metric per ROI and clinical category* ∼ *1 + **Sex** + Age*

The model assessed differences in regional WMH burden or ratio between sexes (variable of interest) for each clinical category, while correcting for age. Linear models and multiple comparison corrections were applied within the CCNA COMPASS-ND cohort only.

#### 2.3.2 Analyses for Aim 4: association between AT-specific WMH burden and cognition

For Aim 4, we used cognitive assessments on a subset of our CCNA COMPASS-ND diagnostic groups, i.e., CU, SCD, MCI, AD, and MCI and AD with high vascular brain injury (N=756; Fig. 5a). Cognitive performance was operationalized as processing speed and cognitive screening using four psychometric tests: Simple Reaction Time (SRT), Choice Reaction Time (CRT), Digit Symbol Substitution Test (DSST), and Montreal Cognitive Assessment (MoCA) (Fig. 5b). SRT, CRT, and DSST evaluate processing speed, which is often one of the earliest cognitive functions to decline with age. The MoCA provides a broader screening of multiple cognitive domains, including executive function, memory, language, and visuospatial ability.

Statistical analyses involved a series of linear regression models, with cognitive performance as the dependent variable and region-size-normalized AT-WMH burden as the independent variable, adjusting for age and sex. A 5% False Discovery Rate threshold was applied to correct for multiple comparisons.

*Aim 4.1. Cognition* ∼ *1 + **WMH burden per ROI** + Clinical category + Sex + Age*

Model assesses how the WMH burden (variable of interest) in specific brain regions is associated with cognitive performance (dependent variable), while adjusting for age, sex, and applying multiple comparison corrections. This analysis was conducted for each cognitive test and adjusting for diagnosis.

*Aim 4.2. Cognition* ∼ *1 + WMH burden per ROI and clinical category + Sex + Age*

Model assesses how the WMH burden (variable of interest) in specific brain regions influences cognitive performance, while adjusting for age, sex, and applying multiple comparison corrections. This analysis was conducted for each cognitive test and diagnosis.

## 3. RESULTS

### 3.1 Cohort Demographics

#### CCNA COMPASS-ND cohort

Brain-size normalized WMH burden values (in cubic centimeters, cm^3^) per clinical category in each ROI are summarized in demographic Table 1.1 for the CCNA COMPASS-ND cohort. Participant ages ranged from 50 to 90 years old, with FTD being the youngest group and AD+V the oldest (Tables 1.1). Whole-brain WMH burden means ranged from 8.53±6.23 (CU) to 31.97±17.43 (AD+V) cm^3^ in CCNA COMPASS-ND participants.

**Table 1.1.** Demographic characteristics and WMH volumes (brain-size normalized and adjusted for AT-size values in cubic centimeters) for the CCNA COMPASS-ND participants used in this study, per clinical category. Participants from a total of ten clinical categories were included.

| Diagnosis | N |  |  |  | WMH burden per region of interest (cm<sup>3</sup>) |  |  |  |  |  |  |  |  |  |
| --- | --- | --- | --- | --- | --- | --- | --- | --- | --- | --- | --- | --- | --- | --- |
|  | Total | F | M | Age | Total<br>M ± SD | ACA<br>M ± SD | MCA<br>M ± SD | PCA<br>M ± SD | LACA<br>M ± SD | LMCA<br>M ± SD | LPCA<br>M ± SD | RACA<br>M ± SD | RMCA<br>M ± SD | RPCA<br>M ± SD |
| CU | 107 | 81 | 26 | 70 ± 6.11 | 8.53 ± 6.23 | 0.82 ± 0.78 | 1.24 ± 0.96 | 0.93 ± 0.56 | 0.39 ± 0.39 | 0.67 ± 0.51 | 0.47 ± 0.30 | 0.43 ± 0.40 | 0.57 ± 0.46 | 0.46 ± 0.28 |
| SCD | 133 | 97 | 36 | 70.4 ± 5.98 | 10.79 ± 9.28 | 1.14 ± 1.37 | 1.62 ± 1.40 | 0.90 ± 0.49 | 0.53 ± 0.63 | 0.86 ± 0.74 | 0.43 ± 0.24 | 0.61 ± 0.76 | 0.76 ± 0.67 | 0.47 ± 0.29 |
| MCI | 240 | 99 | 141 | 72.1 ± 6.69 | 10.55 ± 8.20 | 1.05 ± 1.19 | 1.54 ± 1.25 | 1.10 ± 0.55 | 0.49 ± 0.58 | 0.81 ± 0.66 | 0.52 ± 0.27 | 0.56 ± 0.64 | 0.73 ± 0.61 | 0.58 ± 0.33 |
| MCI+V | 133 | 59 | 74 | 76.5 ± 6.3 | 29.94 ± 17.97 | 3.81 ± 3.07 | 4.45 ± 2.58 | 1.81 ± 0.90 | 1.77 ± 1.42 | 2.32 ± 1.33 | 0.85 ± 0.45 | 2.04 ± 1.69 | 2.12 ± 1.29 | 0.96 ± 0.51 |
| AD | 93 | 40 | 53 | 73.1 ± 7.79 | 14.02 ± 10.14 | 1.52 ± 1.83 | 1.98 ± 1.36 | 1.50 ± 0.73 | 0.73 ± 0.90 | 1.04 ± 0.73 | 0.71 ± 0.39 | 0.80 ± 0.96 | 0.94 ± 0.67 | 0.79 ± 0.40 |
| AD+V | 46 | 20 | 26 | 78.7 ± 5.59 | 31.97 ± 17.43 | 4.22 ± 3.20 | 4.60 ± 2.34 | 2.14 ± 0.93 | 2.05 ± 1.60 | 2.46 ± 1.23 | 1.10 ± 0.59 | 2.17 ± 1.65 | 2.14 ± 1.17 | 1.04 ± 0.44 |
| LBD | 26 | 4 | 22 | 73.3 ± 7.91 | 18.87 ± 14.15 | 2.20 ± 2.00 | 2.70 ± 2.07 | 1.70 ± 1.00 | 1.03 ± 0.92 | 1.39 ± 1.01 | 0.81 ± 0.44 | 1.17 ± 1.15 | 1.31 ± 1.09 | 0.89 ± 0.62 |
| FTD | 23 | 11 | 12 | 65.6 ± 8.89 | 17.39 ± 11.44 | 2.04 ± 1.84 | 2.49 ± 1.58 | 1.53 ± 0.81 | 0.92 ± 0.91 | 1.27 ± 0.86 | 0.82 ± 0.47 | 1.12 ± 0.96 | 1.22 ± 0.82 | 0.71 ± 0.40 |
| PD | 77 | 36 | 41 | 66.8 ± 7.2 | 9.91 ± 9.45 | 0.96 ± 1.12 | 1.51 ± 1.54 | 0.91 ± 0.57 | 0.47 ± 0.52 | 0.81 ± 0.83 | 0.44 ± 0.26 | 0.49 ± 0.61 | 0.70 ± 0.73 | 0.47 ± 0.36 |
| PDD | 49 | 7 | 42 | 72 ± 7.64 | 16.63 ± 12.80 | 1.76 ± 1.60 | 2.47 ± 2.03 | 1.49 ± 0.74 | 0.88 ± 0.88 | 1.34 ± 1.13 | 0.68 ± 0.37 | 0.88 ± 0.79 | 1.13 ± 0.93 | 0.81 ± 0.41 |

#### NIFD cohort

Brain-size normalized WMH burden values (in cubic centimeters, cm^3^) per clinical category in each ROI are summarized in demographic Table 1.2 for the NIFD cohort. Participant ages ranged from 45 to 90 years old. Given differences in imaging parameters, whole-brain WMH burden means were lower for categories in the NIFD cohort: CU_NIFD (1.6±2.8 cm³) and FTD_NIFD (2.5±2.9 cm³).

**Table 1.2.** Demographic characteristics and WMH volumes (brain-size normalized and adjusted for AT-size values in cubic centimeters) for the NIFD participants used in this study, per clinical category. Participants from a total of two clinical categories were included.

| Diagnosis | N |  |  |  | WMH burden per region of interest (cm<sup>3</sup>) |  |  |  |  |  |  |  |  |  |
| --- | --- | --- | --- | --- | --- | --- | --- | --- | --- | --- | --- | --- | --- | --- |
|  | Total | F | M | Age | Total<br>M ± SD | ACA<br>M ± SD | MCA<br>M ± SD | PCA<br>M ± SD | LACA<br>M ± SD | LMCA<br>M ± SD | LPCA<br>M ± SD | RACA<br>M ± SD | RMCA<br>M ± SD | RPCA<br>M ± SD |
| CU | 96 | 53 | 43 | 63.2 ± 7.11 | 1.5993 ± 2.8365 | 0.5463 ± 1.064 | 0.8276 ± 1.6641 | 0.2254 ± 0.2956 | 0.2377 ± 0.5196 | 0.4135 ± 0.8588 | 0.1088 ± 0.1793 | 0.3086 ± 0.5752 | 0.4141 ± 0.8316 | 0.1167 ± 0.1322 |
| FTD | 98 | 46 | 52 | 62.7 ± 7.34 | 2.4739 ± 2.8646 | 1.1497 ± 1.6185 | 1.0029 ± 1.2041 | 0.3214 ± 0.3129 | 0.5983 ± 0.8664 | 0.5186 ± 0.6426 | 0.1527 ± 0.1712 | 0.5513 ± 0.7776 | 0.4843 ± 0.5949 | 0.1686 ± 0.1995 |

### 3.2 Pairwise Comparisons Across Arterial-territories (Aim 1)

#### 3.2.1 AT-specific WMH burden findings – CCNA COMPASS-ND cohort

All clinical categories exhibited a higher WMH burden in the MCA_t_ when compared to ACA_t_ and PCA_t_, despite correcting for AT size differences. However, when comparing the ACA_t_ to the PCA_t_, our analysis unveiled distinct patterns of WMH burden across clinical categories. Specifically, CU, MCI, AD, and PD displayed a higher WMH burden in PCA_t_ compared to ACA_t_. Clinical categories characterized by high vascular brain injury (i.e., MCI+V and AD+V) demonstrated a higher WMH burden in ACA_t_ compared to PCA_t_ (Fig. 2).

**Fig. 2.**
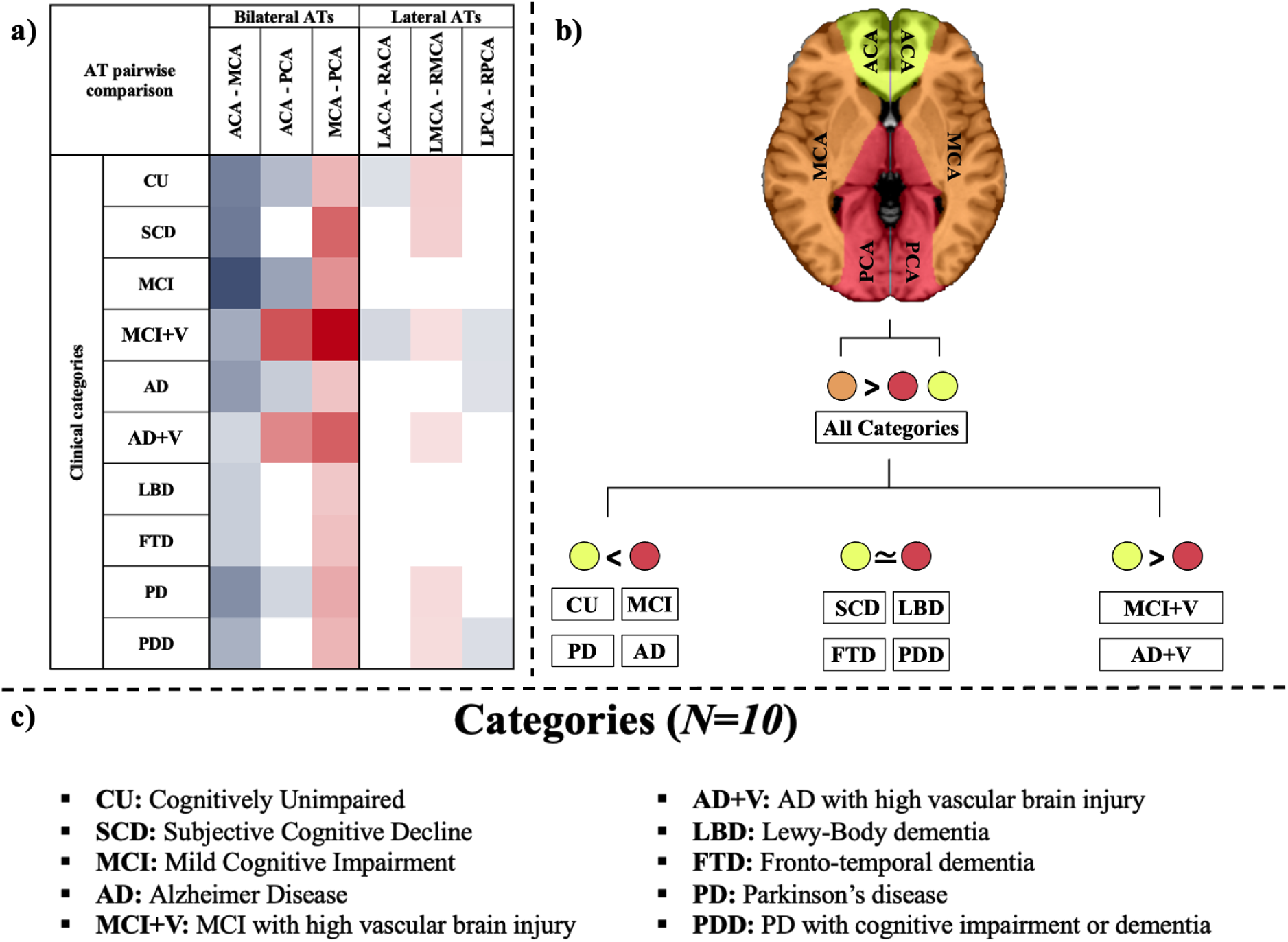
AT-specific WMH comparisons between regions. **a)** Columns indicate pairs of contrasted regions (AT 1 - AT 2), each row referring to a distinct clinical category. The <u>red</u> color represents regions where AT 1 has a significantly *higher* WMH burden than AT 2, with the intensity representing *t-*values. Similarly, the <u>blue</u> color represents regions where AT 1 has a significantly *lower* WMH-metric than AT 2. White cells represent contrasts that exhibit no significant statistical difference. Results are corrected for multiple comparisons using FDR. **b)** Altered visualization for the bilateral ATs section of Fig. 2a, where each color represents a different AT. The mathematical comparative signs indicate the differences in WMH burden between these ATs for each clinical group. **c)** Acronym definition for the ten presented CCNA COMPASS-ND categories.

Left and right asymmetry analyses revealed that most categories (6/10, CU, SCD, MCI+V, AD+V, PD, PDD) demonstrated AT-specific asymmetry in the MCA_t_, with a higher WMH burden on the left side. Additionally, MCI+V and CU had a higher WMH burden in the RACA_t_ and MCI+V, AD, and PDD had a higher WMH burden in the RPCA_t_ (Fig. 2).

##### SCD

Note that we report on SCD findings separately, given that this group differs from controls only due to subjective self-reported cognitive impairment, with no objective impairment detected in cognitive testing. Similar to the other clinical categories, SCD demonstrated a higher WMH burden in the MCA_t_ relative to both the ACA_t_ and PCA_t_ (Fig. 2). However, no significant difference was detected between the ACA_t_ and PCA_t_ (Fig. 2). Within the MCA_t_, an asymmetry in burden distribution was observed, with the LMCA_t_ displaying a higher WMH burden than the RMCA_t_.

#### 3.2.2 AT-specific WMH burden findings – NIFD cohort

Consistent with our FTD findings in the CCNA COMPASS-ND cohort, FTD_NIFD had a higher WMH burden in the MCA_t_ compared to the other ATs. However, FTD_NIFD had a higher WMH burden in the ACA_t_ than in the PCA_t_ (Supplemental Fig. 1), while no significant difference between the ACA_t_ and PCA_t_ was detected in FTD (CCNA COMPASS-ND). Consistent with our CCNA COMPASS-ND findings, the FTD_NIFD cohort showed no asymmetry.

### 3.3 Pairwise Comparisons Across Clinical Categories (Aim 2) (WMH burden)

#### 3.3.1 WMH burden findings – CCNA COMPASS-ND cohort

The results of pairwise group comparisons for whole-brain (or total) and regional WMH burden are summarized in Fig. 3a. The number of categories for which a given group had a higher WMH burden in a specific region are indicated in Fig. 3b. These values are alternatively visualized in the frequency histogram shown in Fig. 3c.

**Fig. 3.**
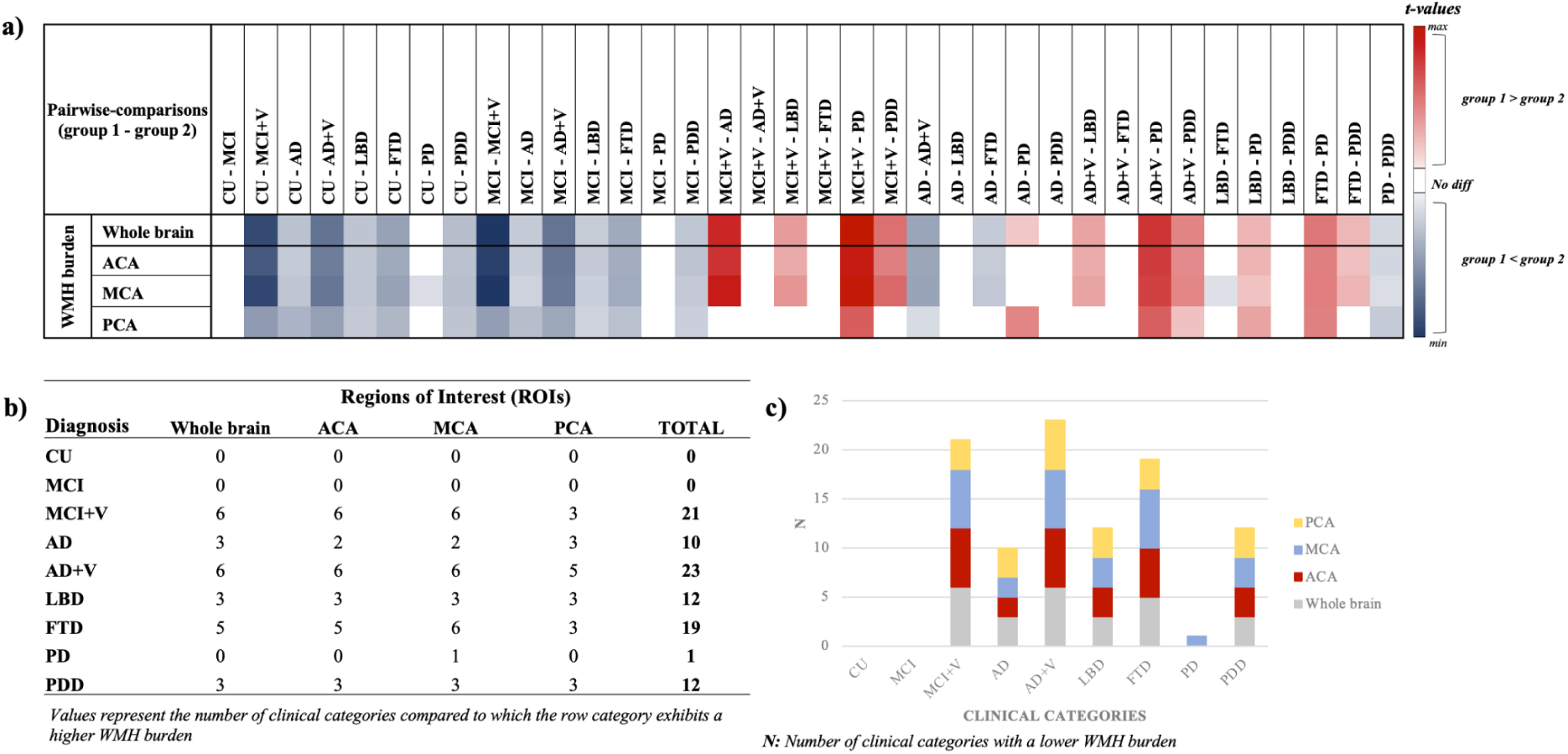
AT-specific WMH burden comparisons between clinical categories. **a)** Columns indicate pairs of contrasted groups (group 1 - group 2), each row referring to a distinct region. The red color represents regions where group 1 has a significantly higher WMH burden than group 2, with the gradient representing *t-*values. Similarly, the blue color represents regions where group 1 has a significantly lower WMH burden than group 2. White cells represent contrasts that exhibit no significant statistical difference. Results are corrected for multiple comparisons using FDR. **b)** Figure summarizes group comparison findings in Fig. 3a. Rows represent specific groups, columns represent ATs, and values indicate the frequency of each group having a higher WMH burden compared to others. **c)** Histogram visualization of findings in Fig. 3b.

##### Whole-brain findings

Pairwise comparisons between clinical categories found those characterized by a high vascular brain injury (i.e., MCI+V and AD+V) to have a significantly higher whole-brain WMH burden compared to all non-vascular clinical categories, except FTD (Fig. 3). In the non-vascular clinical categories, FTD had a significantly higher whole-brain WMH burden, except when compared to LBD. Of the remaining six clinical categories, LBD, AD, and PDD had a significantly higher whole-brain WMH burden compared to PD, MCI, and CU (Fig. 3).

##### AT-specific findings

In the ACA_t_ and MCA_t_, WMH burden pairwise comparisons between clinical categories were consistent with whole-brain findings, except for AD, which was not significantly different from PD (Fig. 3). Pairwise comparisons in the MCA_t_ showed two additional group differences in WMH burden that were not detected when looking at the whole-brain, namely, FTD higher than LBD and PD higher than CU (Fig. 3). Pairwise comparisons between clinical categories in the PCA_t_ showed fewer group differences compared to whole-brain findings, with no significant difference detected between (i) MCI+V versus AD, LBD, or PDD, (ii) FTD versus AD or PDD, and (iii) AD+V versus LBD (Fig. 3).

#### SCD

##### Whole-brain findings

Pairwise comparisons between SCD and other clinical categories showed that SCD exhibited a whole-brain WMH burden that was (i) higher compared to CU and (ii) lower compared to high vascular injury burden categories (MCI+V and AD+V), and dementia categories (AD, LBD, FTD, and PDD) (see Supplemental Fig. 2).

##### AT-specific findings

In the ACA_t_ and MCA_t_, WMH burden pairwise comparisons between SCD and other clinical categories were consistent with whole-brain findings, except when compared to AD (see Supplemental Fig. 2). Moreover, SCD had a lower WMH burden in the PCA_t_ compared to MCI and AD.

#### 3.3.2 WMH burden findings – NIFD cohort

When comparing CU_NIFD and FTD_NIFD cohorts, our WMH burden findings mostly reproduced our CCNA COMPASS-ND CU-FTD comparative findings (i.e., higher WMH burden in FTD across all bilateral and lateral ATs compared to CU). The only exception was the RPCA_t_ WMH burden, where no statistically significant difference was detected between CU_NIFD and FTD_NIFD groups (Supplemental Fig. 1).

#### 3.3.3 WMH ratio findings – CCNA COMPASS-ND cohort

The results of pairwise group comparisons for regional WMH ratio are summarized in Fig. 4a. The number of categories for which a given group had a higher WMH ratio in a specific region are indicated in Fig. 4b. These values are alternatively visualized in the frequency histogram shown in Fig. 4c.

**Fig. 4.**
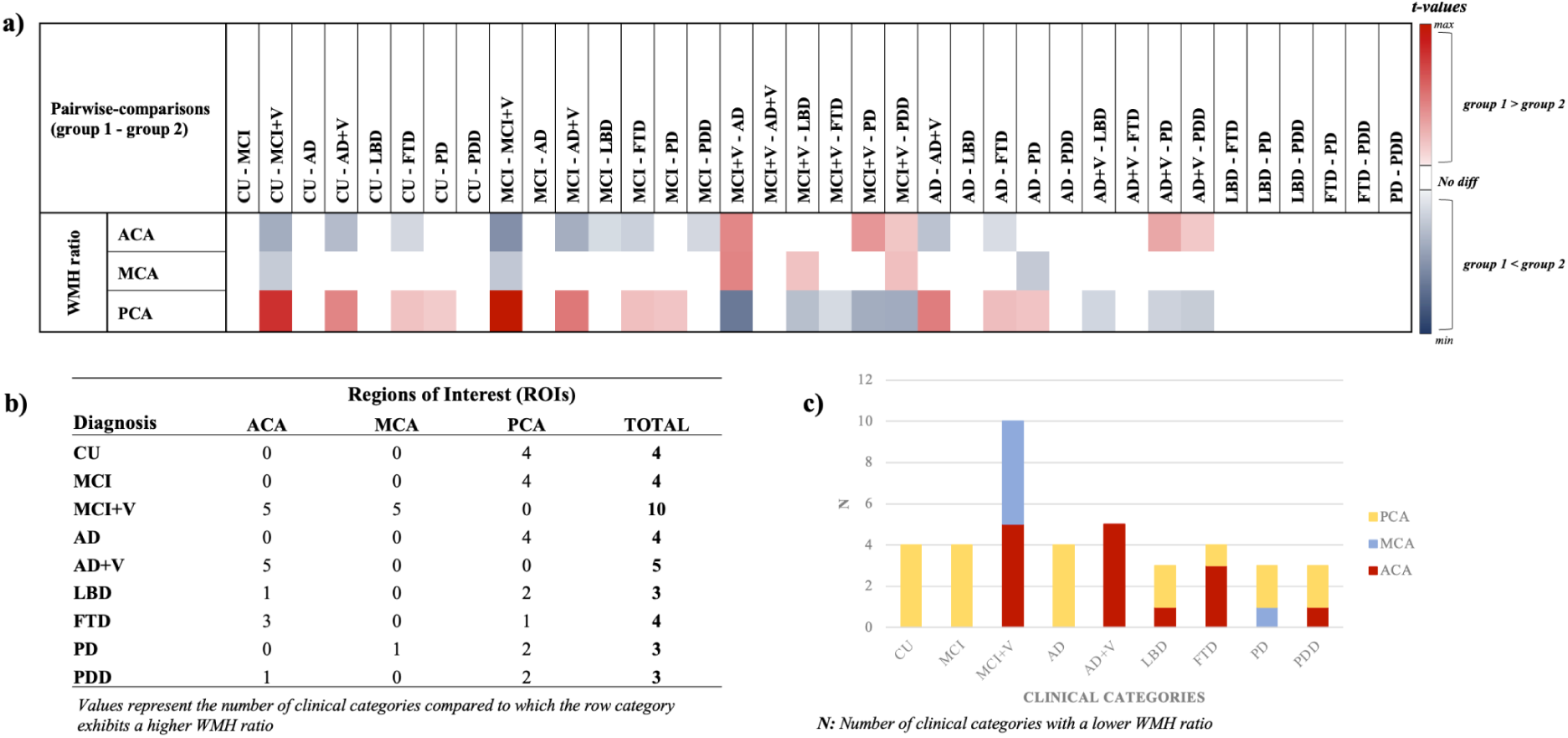
AT-specific WMH ratio comparisons between clinical categories. **a)** Columns indicate pairs of contrasted groups (group 1 - group 2), each row referring to a distinct AT. The red color represents regions where group 1 has a significantly higher WMH ratio than group 2, with the gradient representing t-values. Similarly, the blue color represents regions where group 1 has a significantly lower WMH ratio than group 2. White cells represent contrasts that exhibit no significant statistical difference. Results are corrected for multiple comparisons using FDR. **b)** Figure summarizes group comparison findings in Fig. 4a. Rows represent specific groups, columns represent ATs, and values indicate the frequency of each group having a higher WMH burden compared to others. **c)** Histogram visualization of findings in Fig. 4b.

WMH ratio pairwise comparisons between clinical categories (N=45) showed that most group differences were detected in the PCA_t_ (N=19), followed by the ACA_t_ (N=15) and the MCA_t_ (N=6) (Fig. 4). Our findings highlighted a contrasting relationship between the ACA_t_ and PCA_t_: in most cases (N=13; 62%), when the WMH ratio was lower in the ACA_t_ it was higher in the PCA_t_, and vice-versa (Fig. 4).

Categories with a high vascular brain injury (i.e., MCI+V and AD+V) had significantly (i) higher WMH ratio in the ACA_t_ compared to all other categories, except for LBD and FTD where no difference was detected, and (ii) lower WMH ratio in the PCA_t_ compared to all other categories, except for FTD where no difference was detected when compared to AD+V (Fig. 4). Only MCI+V had a higher WMH ratio in the MCA_t_ than all categories, except for FTD and PD. As for the remaining categories, FTD had a WMH ratio that was higher in the ACA_t_ and lower in the PCA_t_ compared to AD, MCI, and CU (Fig. 4). PD had a WMH ratio that was (i) lower in the PCA_t_ compared to AD, MCI, and CU (similar to FTD), and (ii) higher in the MCA_t_ compared to AD (Fig. 4). Finally, MCI had a higher WMH ratio than LBD, FTD, and PDD in the ACA_t_.

##### SCD

The results of pairwise group comparisons for regional WMH ratio are summarized in Supplemental Fig. 3. Compared to categories with a high vascular injury burden, SCD had a lower ACA_t_ WMH ratio and a higher PCA_t_ WMH ratio. SCD had a higher MCA_t_ WMH ratio and a lower PCA_t_ WMH ratio than CU, MCI, and AD. Lastly, SCD had a higher ACA_t_ WMH ratio than MCI.

#### 3.3.4 WMH ratio findings – NIFD cohort

When comparing CU_NIFD and FTD_NIFD cohorts, our WMH ratio findings when comparing CU and FTD (CCNA COMPASS-ND) were not reproduced (i.e., higher WMH ratio than CU in the ACA_t_ and lower in the PCA_t_). The NIFD cohort showed no statistically significant WMH ratio differences between CU_NIFD and FTD_NIFD (Supplemental Fig. 1).

### 3.4 Sex Analysis *–* WMH burden and Ratio (Aim 3)

Compared to women, men exhibited higher whole-brain and MCA_t_ WMH burden in CU, MCI+V, and FTD. Additionally, men showed a higher ACA_t_ WMH burden than women in MCI+V, as well as a higher PCA_t_ WMH burden in MCI, FTD, and PD (Table 2.1). In terms of the WMH ratio analyses, women exhibited a higher ACA_t_ WMH ratio in SCD and MCA_t_ and a lower MCA_t_ WMH ratio in PDD (Table 2.2).

**Table 2.1.**
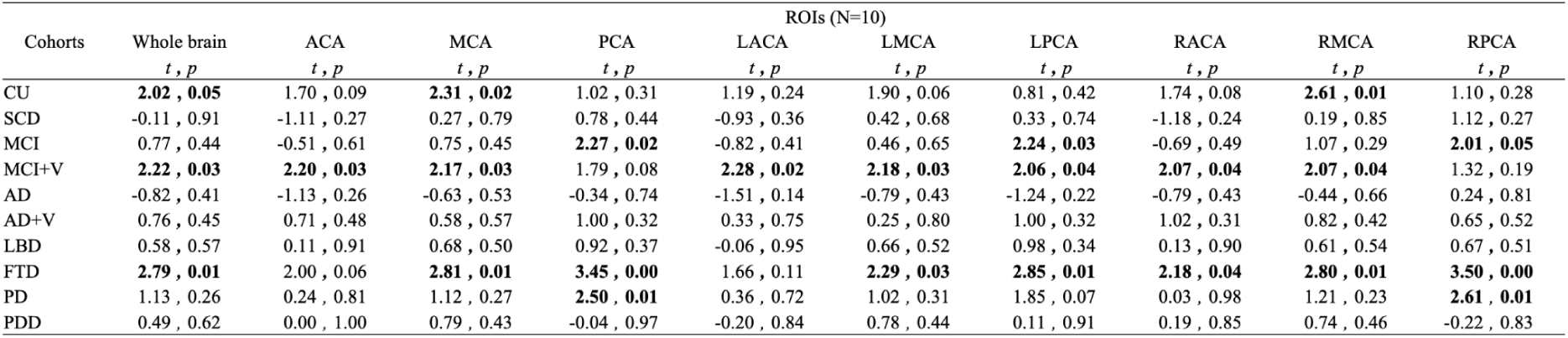
WMH burden Sex-analyses. Negative *t*-values represent instances where men had a lower WMH burden, and vice versa for positive *t*-values. Statistically significant findings after FDR correction are shown in bold.

**Table 2.2.** WMH ratio Sex-analyses. Negative *t*-values represent instances where men had a lower WMH ratio, and vice versa for positive *t*-values. Statistically significant findings after FDR correction are shown in bold.

| Cohorts | ROIs (N=10) |  |  |  |  |  |  |  |  |
| --- | --- | --- | --- | --- | --- | --- | --- | --- | --- |
|  | ACA<br>*t, p* | MCA<br>*t, p* | PCA<br>*t, p* | LACA<br>*t, p* | LMCA<br>*t, p* | LPCA<br>*t, p* | RACA<br>*t, p* | RMCA<br>*t, p* | RPCA<br>*t, p* |
| CU | 0.39, 0.69 | 1.49, 0.14 | -1.25, 0.21 | -0.40, 0.69 | 0.33, 0.74 | -1.29, 0.20 | 0.54, 0.59 | **2.06, 0.04** | -0.88, 0.38 |
| SCD | **-2.48, 0.01** | 1.36, 0.18 | 1.27, 0.21 | -1.51, 0.13 | 1.33, 0.19 | 0.61, 0.54 | **-2.20, 0.03** | 0.84, 0.40 | 1.65, 0.10 |
| MCI | **-2.24, 0.03** | -0.75, 0.45 | 1.85, 0.07 | **-2.65, 0.01** | -1.30, 0.20 | 1.76, 0.08 | -1.17, 0.24 | 0.29, 0.78 | 1.45, 0.15 |
| MCI+V | 1.65, 0.10 | -0.28, 0.78 | -0.88, 0.38 | 1.73, 0.09 | -0.12, 0.91 | -0.41, 0.69 | 1.22, 0.22 | -0.29, 0.77 | -1.10, 0.27 |
| AD | -1.27, 0.21 | 0.56, 0.58 | 0.81, 0.42 | -1.83, 0.07 | 0.00, 1.00 | -0.65, 0.52 | -0.46, 0.65 | 0.85, 0.39 | 1.64, 0.11 |
| AD+V | 0.26, 0.80 | -1.09, 0.28 | 0.05, 0.96 | -0.57, 0.57 | -1.80, 0.08 | 0.28, 0.78 | 0.88, 0.38 | -0.09, 0.93 | -0.32, 0.75 |
| LBD | -1.03, 0.31 | 0.66, 0.52 | 0.55, 0.59 | -1.00, 0.33 | 0.51, 0.62 | 0.32, 0.75 | -0.78, 0.44 | 0.29, 0.78 | 0.35, 0.73 |
| FTD | -0.19, 0.85 | -0.14, 0.89 | 0.08, 0.94 | -0.32, 0.75 | -0.90, 0.38 | -0.08, 0.94 | 0.14, 0.89 | 0.26, 0.80 | 0.23, 0.82 |
| PD | -1.93, 0.06 | -0.27, 0.79 | 1.36, 0.18 | -1.38, 0.17 | -0.44, 0.66 | 0.49, 0.63 | -1.86, 0.07 | 0.15, 0.88 | 1.62, 0.11 |
| PDD | -0.81, 0.42 | **2.36, 0.02** | -0.64, 0.52 | -1.25, 0.22 | 1.86, 0.07 | -0.38, 0.71 | -0.11, 0.91 | 1.72, 0.09 | -0.84, 0.41 |

### 3.5 Association Between AT-specific WMH burden and Cognition (Aim 4)

#### 3.5.1 Analysis independent of diagnosis (Aim 4.1)

Our results revealed that, when adjusting for clinical diagnosis: (i) SRT scores were associated with ACA WMH burden (*p*=0.002; *t*=2.44), (ii) CRT and MoCA scores were associated with PCA WMH burden (CRT: *p*=0.002; *t*=2.40 | MoCA: *p*=0.001; *t*=-3.21), and (iii) DSST scores were associated with WMH burden across all ATs (PCA: *p*<0.001; *t*=-3.96 | MCA: *p*=0.004; *t*=-2.83 | ACA: *p*=0.02; *t*=-2.26).

#### 3.5.2 NDD-specific analysis (Aim 4.2)

MCI and AD were the only categories to exhibit AT-specific WMH burden associations with cognitive performance (Fig. 5c). In MCI, lower DSST scores were associated with higher PCA WMH burden (*p*=0.02; *t*=-2.78) (Fig. 5d). In AD, a lower percentage of correct answers in CRT tasks was associated with higher ACA (*p*=0.006; *t*=-3.32) and MCA (*p*=0.02; *t*=-2.59) WMH burden (Fig. 5e).

**Fig. 5.**
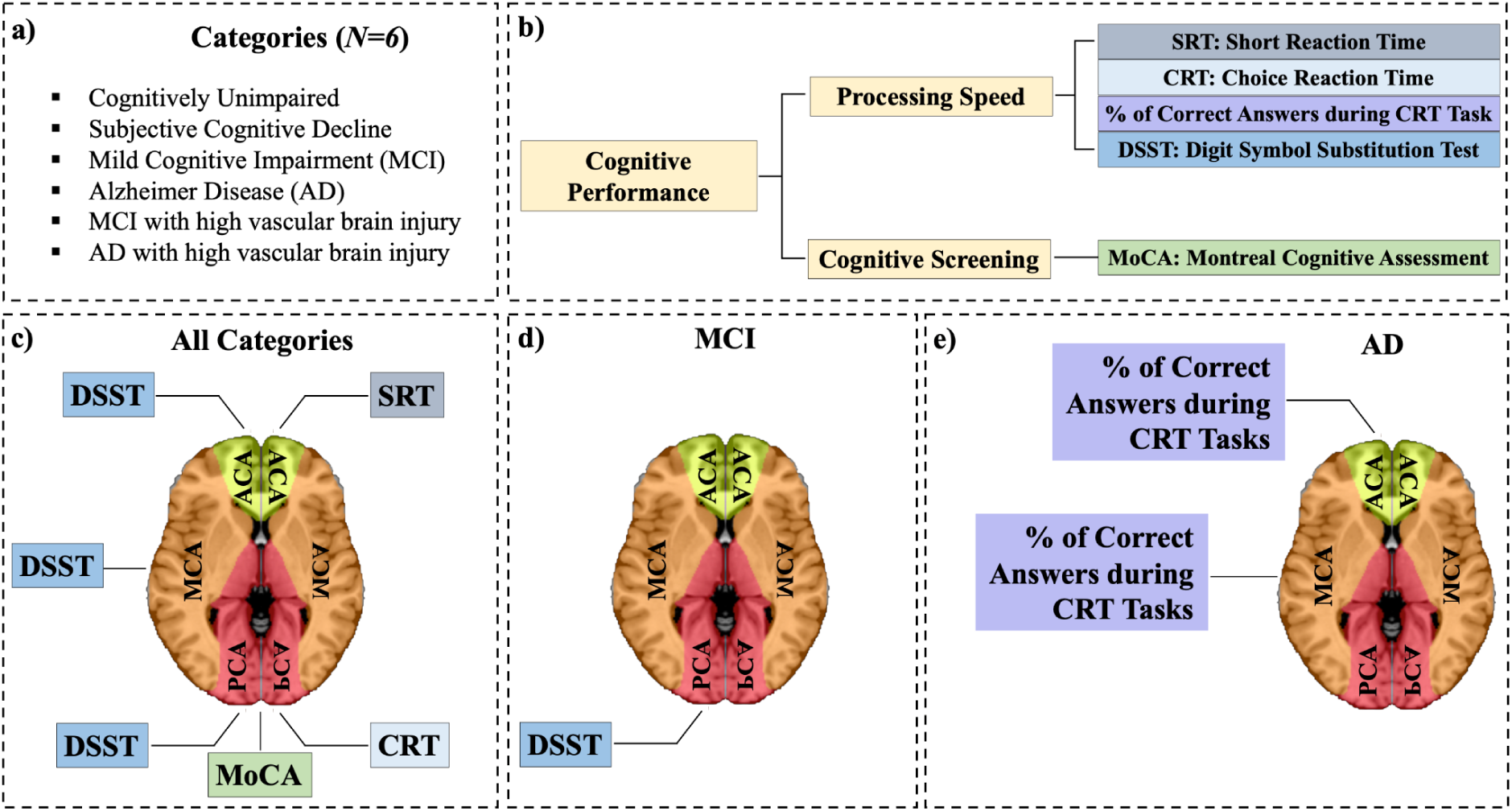
Associations between AT-specific WMH burden and Cognitive Performance. **a)** Clinical categories analyzed. **b)** Tasks used for cognitive assessment. **c)** Across categories analysis: AT-specific WMH burden associations with cognitive scores from different cognitive assessments. **d)** NDD-specific analysis: PCA WMH burden was associated with DSST scores in MCI. **e)** NDD-specific analysis: ACA and MCA WMH burden were significantly associated with the percentage of Correct Answers in CRT Tasks in AD. Note: All presented results maintain significance (*p*<0.05) after correcting for multiple comparisons.

## 4. DISCUSSION

In this study, we mapped WMH across ATs associated with the three major cerebral arteries (ACA, MCA, PCA) in both hemispheres in ten clinical categories, uncovering unique AT-specific WMH signatures in certain NDDs and their prodromes. These major cerebral arteries originate from the Circle of Willis [64], with the ACA and MCA supplied by the anterior circulation and the PCA by the posterior circulation. They serve distinct brain regions: the ACA irrigates the medial hemispheres and superior frontal/parietal lobes; the MCA covers the inferior frontal, inferolateral parietal, and lateral temporal lobes; and the PCA supplies the occipital and medial temporal lobes [65]. Pathological changes in these arteries can lead to AT-specific disturbances and increased WMH burden [31,32].

### 4.1 Insights From WMH burden Distributions Across and Within ATs

#### 4.1.1 Higher MCA_t_ vulnerability and distinct AT patterns across clinical categories

Our transdiagnostic investigation of WMH burden differences across bilateral ATs (MCA_t_, ACA_t_, PCA_t_) led to two main conclusions.

First, we found that the MCA_t_ demonstrated the highest WMH burden across all clinical categories assessed, even after adjusting for territory size. Consistent with our finding, the MCA_t_ has been reported to have a higher burden of cerebrovascular pathologies compared to other ATs, including infarcts [46,66–70], atherosclerosis [71,72] and WMH [46]. This may be because the MCA is the cerebral artery most frequently affected by pathology [73,74], partly due to the higher hemodynamic-stress associated with its geometry (e.g., diameter, bifurcation angle) and other attributes [75,76]. An example of the latter is that the MCA exhibits the highest sensitivity to blood pressure variations among the anterior circulation arteries [77], an established risk--factor for WMH and other cerebrovascular pathologies [77,78].

Second, our ACA_t_-PCA_t_ comparisons revealed AT-specific WMH burden patterns unique to clinical categories with similar whole-brain WMH burden. Specifically, categories with whole-brain WMH volumes of <15 cm³ (CU, AD continuum, PD) exhibited higher WMH burden in the PCA_t_ compared to the ACA_t_. Conversely, in clinically recognized high vascular injury categories, namely MCI+V and AD+V with whole-brain WMH volumes of >29 cm³, the WMH burden was higher in the ACA_t_ than in the PCA_t_.

Supporting our ACA_t_-PCA_t_ findings in the AD continuum, studies have consistently reported an increased posterior WMH burden in these categories [11,43,79–82]. This posterior predominance of WMH in AD is thought to stem from a combination of amyloid pathology [43,83–85] and cerebrovascular issues [86–88]. The main mechanisms include (i) amyloidosis-associated neuroinflammation commencing in the posterior white matter [89,90], (ii) increasing cerebral amyloid angiopathy-associated ischemic injury predominantly affecting posterior brain vessels, partly due to differential aging of the posterior circulation (e.g., increased collagen content) [35,83,91–94], and (iii) higher frequency of severe PCA occlusion by atherosclerotic lesions [95]. Compared to AD, there are relatively few studies elucidating our ACA_t_-PCA_t_ findings in CU and PD. In CU, our findings are supported by a large sample-size study, showing that posterior WMH burden increased more rapidly than in anterior regions [96]. In PD, vascular pathology is increasingly recognized as a contributor to disease pathophysiology, potentially sustaining or exacerbating neuronal degeneration in posterior regions [97]. Additionally, the substantia nigra, which is particularly affected in PD [98,99] is vascularized by the posterior circulation through the PCA and basilar artery [99].

Supporting our ACA_t_-PCA_t_ findings, the increased anterior WMH burden observed in the high vascular injury categories (MCI+V, AD+V) aligns with studies reporting that individuals with vascular cognitive impairment and dementia have a higher proportion of the total WMH burden accumulating in the anterior relative to posterior regions [100]. Studies also indicate that (i) more vascular risk-factors (e.g., age, hypertension) are associated with anterior (e.g., ACA_t_ [46]) than with posterior WMH burden [11,46,80,85], and (ii) early myelin changes in small vessel disease initially involve the frontal lobe, followed by gradual involvement of the parietal, temporal, and occipital lobes [101].

Among the two FTD cohorts investigated, the NIFD cohort exhibited a higher WMH burden in the ACA_t_ compared to the PCA_t_, although this difference was not significant in the smaller CCNA cohort. While FTD mostly affects both the frontal and temporal lobes [102–104], our findings align with previous research indicating greater hypoperfusion in the frontal compared to the temporal lobe in FTD [105]. This is particularly relevant as the frontal lobe is largely supplied by the ACA [106], which may explain the observed higher WMH burden in the ACA_t_. Notably, progranulin mutations, present in 10-20% of the FTD population [107,108] and known to exacerbate WMH burden [109], are primarily linked to glial dysfunction [110]. However, growing evidence suggests that cerebrovascular factors may also play a role in WMH development in both genetic and sporadic forms of the disease. This is supported by findings that (i) progranulin influences vascular tone [111,112], and that progranulin-deficient mice exhibit microvascular alterations in the brain [113] and (ii) hypoperfusion in sporadic FTD leads to the degeneration of a subset of astrocytes [114].

#### 4.1.2 Left-sided hemispheric dominance in WMH burden across clinical categories

Our transdiagnostic investigation of WMH burden differences within ATs identified hemispheric asymmetries in seven clinical categories (CU, SCD, MCI+V, AD, AD+V, PD, PDD), with 86% showing higher WMH burden in the LMCA_t_. Hemispheric asymmetries in cerebrovascular properties (e.g., morphology, hemodynamics) contribute to the regional occurrence of vascular pathologies [115], which can increase the ipsilateral WMH burden [116,117]. Emerging literature indicates that the MCA exhibits strong left-sided arterial dominance from the neonatal stage, based on metrics of geometry and hemodynamics [115,118,119]. This early asymmetric blood flow weakens LMCA vascular endothelial functions [115], contributing to the prevalence of left-sided MCA injuries [115,118–120], such as the higher WMH burden we observed. NDD-specific brain hemispheric asymmetry may also play an additive or independent role [121].

### 4.2 Insights From AT-specific WMH metrics Across Clinical Categories

#### 4.2.1 Advantage of using both burden and ratio metrics

We found that the majority of WMH burden comparisons (36/45; 80%) between clinical categories showed findings in the ATs that followed the whole-brain pattern – for example, compared to MCI, MCI+V had a higher WMH burden in the whole-brain, as well as in all three ATs. Adjusting for the whole-brain WMH burden (i.e., WMH ratio metric) allowed us to uncover additional characteristics of WMH distribution across ATs per clinical category, independent of overall WMH burden. For example, compared to MCI, MCI+V had a higher ACA_t_ and a lower PCA_t_ WMH ratio.

#### 4.2.2 AT-specific WMH distribution in CU: Insights from cross-clinical comparisons

We found that CU consistently exhibited a lower WMH burden across ATs compared to the dementias (AD, AD+V, FTD, LBD, PDD), although the difference with MCI did not reach statistical significance. Despite the limited AT-specific studies in the literature, our findings align with broader research on whole-brain and regional WMH burden in AD and FTD. Specifically, individuals with AD demonstrated greater WMH accumulation both globally and regionally compared to CU [79,81,122,123]. Our AD+V (and MCI+V) findings align with studies that have established a significant association between WMH increase and mixed dementia [16,124–126]. Additionally, FTD individuals, particularly those with the progranulin mutation, typically have a higher WMH burden than CU [127,128]. Although progranulin status was not available, some of our participants may carry this mutation, as it occurs in 10-20% of the FTD population [107,108]. In contrast to AD and FTD, there exists a lack of consensus in the literature regarding WMH burden in LBD and PDD. Earlier studies reported no significant difference in WMH burden severity relative to CU [122], while recent studies indicate a significantly more severe burden in LBD [129] and PDD [130]. Finally, with regards to PD, although our findings indicated that the increased WMH burden in PD was more pronounced in the MCA_t_, lobar studies reported that PD had greater WMH volume in the occipital region compared to controls [131]. Our results further showed no significant difference in whole-brain WMH burden between CU and PD, consistent with the literature [131].

##### WMH ratio

Most clinical categories (SCD, MCI+V, AD+V, FTD, PD) showed at least one regional WMH ratio difference compared to CU, with a consistently lower WMH ratio in the PCA_t_. This suggests that these categories might accumulate WMH disproportionately across ATs, with relatively less accumulation in the PCA_t_. Further research is needed to understand why WMH tends to accumulate more in the anterior brain regions in these pathologies compared to normal aging.

#### 4.2.3 Proportionate increase in WMH across ATs from MCI to AD

Our pairwise comparisons of WHM burden revealed that MCI individuals had a lower WMH burden compared to those with AD, both in the whole-brain and across all bilateral and lateral ATs (Fig. 3a; Supplemental Fig. 4). The observed AT WMH burden findings in AD are concordant with increased (i) whole-brain WMH burden and (ii) regional WMH burden extracted using other parcellation schemas (e.g., lobar, deep/periventricular), compared to MCI [26,79,81] as well as CU individuals [79,81,123]. The increased WMH burden in the advanced stage of the AD continuum (i.e., dementia stage) was expected given that cerebrovascular perturbations appear early in the disease course (around the same time as Aβ) in both autosomal and sporadic forms of AD, and progress with the severity of the disease [132–136][87]. Moreover, in sporadic AD, vascular abnormalities have been found to be ∼80% more pronounced than other AD biomarkers (e.g., Aβ deposition) from an early stage of the disease, well before clinical onset [135].

While AD had a higher WMH burden than MCI across all ATs, we did not detect any significant WMH ratio findings between the two clinical categories, or when compared to CU. Our observations align with voxel-based literature findings showing that although MCI and AD had a higher WMH burden than CU, this difference disappeared after adjusting for whole-brain WMH burden [123]. Collectively, our findings suggested a proportionate increase in WMH across the ATs as MCI progresses to AD. However, as our findings were based on cross-sectional data, longitudinal studies are needed to confirm these results.

#### 4.2.4 Absence of differences in AT-specific WMH metrics between MCI+V and AD+V

We found no significant difference in WMH metrics (whole-brain or AT-specific) between MCI+V and AD+V. Since their diagnostic criteria did not rely solely on WMH burden [137–139], they may exhibit AT-specific differences in other vascular pathologies [140,141], such as microbleeds and infarcts. Although no significant difference in microbleed count was found between the two categories in our CCNA COMPASS-ND cohort [142], we did not explore their AT-specific distributions, as it fell outside the scope of our study. We recommend that future studies address this aspect.

#### 4.2.5 Distinct AT-specific WMH patterns in vascular versus neurodegenerative conditions

We found that the vascular categories (MCI+V, AD+V) had a higher WMH burden than MCI and AD across all investigated regions, except in the PCA_t_, where MCI+V and AD had similar burden. The latter finding was not unexpected given that, as previously mentioned (Section 4.1), WMH accumulation in posterior brain regions (including the PCA_t_) is common in the AD continuum [42], with the greatest burden likely occurring at the dementia stage. We further found that vascular categories displayed a higher WMH ratio in the ACA_t_ and a lower ratio in the PCA_t_ when compared to MCI and AD. These findings align with literature indicating that (i) individuals with vascular dementia exhibit more anterior than posterior white matter abnormalities compared to AD [100] and (ii) WMH in posterior brain regions may be secondary to AD pathology (i.e., Aβ) [11,85,143,144], whereas WMH in frontal regions are more strongly associated with vascular risk-factors such as hypertension [11,42,46,80,85].

#### 4.2.6 Contrasting AT-specific WMH metrics between FTD and AD

FTD is often misdiagnosed as AD or other conditions including psychiatric disorders [145–148]. Given this, it is important to note that we observed distinct differences in AT-based WMH metrics between FTD and AD. Specifically, FTD exhibited a higher WMH burden in the ACA_t_ and MCA_t_ regions, along with a higher ACA_t_ and lower PCA_t_ WMH ratio. These findings are consistent with the contrasting spatial patterns observed between FTD and AD using other brain imaging-derived measures, such as hypoperfusion, brain atrophy, and perturbed blood-oxygen-level-dependent (BOLD) signal [149–155]. Our results, combined with existing literature, underscored distinct spatial patterns that could help mitigate the frequent misdiagnosis of FTD as AD [145].

#### 4.2.7 LBD and PDD share similar AT-specific WMH patterns, unlike PD

PD, PDD, and LBD are characterized by the presence of abnormal alpha-synuclein deposits in Lewy bodies [156,157]. Our results showed no significant differences in WMH burden or ratio between LBD and PDD across all regions. Both LBD and PDD exhibited a higher WMH burden than PD in the whole-brain and bilateral ATs, though WMH ratios were not statistically different. This aligns with literature indicating a greater WMH burden in PDD compared to PD [158,159].

LBD and PDD showed no significant differences in WMH burden or ratios compared to AD, consistent with reports of no overall WMH differences between AD and LBD [160]. In contrast, PD had a lower total WMH burden than AD, particularly in the PCA_t_, which is often associated with AD-related pathologies (see section 4.1.1).

#### 4.2.8 Distinct AT-specific WMH Patterns in SCD compared to CU and AD continuum

SCD refers to self-reported worsening of cognitive performance or increasing difficulties with memory and thinking [161]. Individuals with SCD have twice the risk of developing MCI or dementia compared to normal aging individuals [162,163]. Identifying objective metrics that differentiate SCD from normal aging is crucial for early detection. Our findings indicated that SCD had a higher whole-brain WMH burden than CU, particularly in the ACA_t_ and MCA_t_, suggesting a distinct WMH profile compared to normal aging. While not all individuals with SCD progress to MCI or AD, longitudinal studies estimate that approximately 20-45% eventually receive such a diagnosis [164–171]. Our results showed that SCD had a significantly lower WMH burden in the PCA_t_ compared to MCI, though this difference was not observed at the whole-brain level. In comparison to AD, SCD had a significantly lower WMH burden in the PCA_t_ and at the whole-brain level, suggesting progressive posterior WMH accumulation in AD.

### 4.3 Sex-specific Differences

Sex-specific differences in WMH burden were observed across several clinical categories (Table 2.1). Consistent with literature suggesting that men tend to experience greater cardiovascular risk factor-related differences in total brain volumes than women [172], men were found to have a higher WMH burden in the whole-brain (controlled for brain size via the use of a standardized brain space) and/or an arterial region across several categories (CU, MCI, MCI+V, FTD, PD). However, after controlling for total WMH burden (i.e., using the WMH ratio), women exhibited a higher ACA_t_ WMH ratio in SCD and MCI (Table 2.2). The literature suggests that in normally aging elderly cohorts, women tended to have a higher proportion of WMH compared to men, relative to the size of their white matter [173–175]. However, this trend may not have been reflected in our findings due to the high proportion of women (80%) in our CU sample, potentially leading to an underrepresentation of WMH distribution in men. Research further reports that women had slower WMH progression with age across the total brain, particularly in the frontal region [176].

### 4.4 Cognition

Cognition results revealed that greater WMH burden was associated with poorer cognitive performance, indicated by higher SRT and CRT scores, as well as lower MoCA scores, DSST scores, and the percentage of correct answers in CRT tasks (see Fig. 5). Mounting evidence suggests that WMH exert an independent effect on cognition in AD, which is additive to the effects of core proteinopathies, such as Aβ and tau [177]. A recent study found that WMH burden in PCA sub-regions contributes to lower cognition, independent of Aβ deposition or atrophy in early AD [11]. Supporting this finding, we observed an association between PCA WMH burden and processing speed at the MCI stage (Fig. 5). However, this association evolved to the ACA and MCA regions at the AD stage (Fig. 5). Since AD is characterized by increased Aβ deposits in areas perfused by the ACA and MCA [178], further investigation is warranted to determine whether the observed associations are driven by WMH or other AD pathologies.

### 4.5 Limitations and Future Directions

Notwithstanding the significant findings, this study is not without its limitations. The ROI-based approach does not allow the detection of regional WMH variations within the ROIs. While it offers advantages in terms of noise robustness and statistical power, considering more fine-grained analyses of WMH could uncover additional insights into their neuropathological characteristics and implications [26]. Another limitation was the large inter-individual variations in AT-specific WMH, which may reduce the reliability and accuracy of the identified NDD signatures at the individual level. Future work exploring the potential of the NDD signatures characterized in this study to accurately diagnose individuals to their respective clinical categories will be essential. By leveraging advanced deep learning techniques, it could be possible to determine the diagnostic accuracy and generalizability of the identified NDD signatures, potentially paving the way for more effective and precise diagnostic tools in the future. Other future directions include studying our AT-specific findings in conjunction with vascular (e.g., blood pulsatility, arterial density, vessel diameter) and non-vascular (amyloid, tau) features in healthy individuals and different dementia typesSuch an approach would provide insights into the pathophysiology of WMH and their role in the NDDs. Future research is warranted to confirm and expand upon our findings.

## 5. CONCLUSION

Our research underscores the importance of assessing WMH distribution through vascular-based brain parcellation, revealing ATs more susceptible to WMH formation across various NDDs. Understanding these WMH patterns within specific ATs is essential due to their alignment with the brain’s vascular architecture and role in NDD pathophysiology. By mapping WMH across ten clinical categories, we provided insights that could help differentiate between disorders. Distinct WMH patterns for several diagnoses suggest that incorporating vascular considerations into imaging criteria may enhance diagnostic precision. While further research is needed, our study lays a strong foundation for integrating vascular insights into diagnostic practices and developing targeted interventions in neurodegeneration.

## Data Availability

All data produced in the present work are contained in the manuscript

## 6. ACKNOWLEDGMENTS

This research was funded by Fonds de Recherche Québec – Santé (FRQS) Chercheurs boursiers Junior 1 (2020–2024) and the Fonds de soutien à la recherche pour les neurosciences du vieillissement from the Fondation Courtois (A.B.). We would also like to thank the following organizations for trainee scholarships: VAST Health Research Training Program MSc Scholarship-2022 and Fondation Lemaire and CIMA-Q MSc Scholarship-2023 (I.H); FRQS bourse de formation à la maîtrise (2021) and FRQS bourse de formation au doctorat (2024) (F.E.D.); VAST Health Research Training Program Doctoral Scholarship-2023 (M.S.).

This research used data from CCNA COMPASS-ND, which was supported by a grant from the Canadian Institutes of Health Research (CIHR), with funding contributions from various partner organizations. We would like to thank Randi Pilon for her help with the CCNA-LORIS database. Finally, we are thankful to have received funding from CCNA Team 9 and the CCNA Women Sex Gender Dementia Program.

This research used data from the Frontotemporal Lobar Degeneration Neuroimaging Initiative (FTLDNI also known as NIFD). Data collection and sharing for this project was funded by the Frontotemporal Lobar Degeneration Neuroimaging Initiative (National Institutes of Health Grant R01 AG032306). The study is coordinated through the University of California, San Francisco, Memory and Aging Center. FTLDNI data are disseminated by the Laboratory for Neuro Imaging at the University of Southern California.The investigators at NIFD/FTLDNI contributed to the design and implementation of FTLDNI and/or provided data, but did not participate in analysis or writing of this report. The FTLDNI investigators included the following individuals: Howard Rosen, University of California, San Francisco (PI); Bradford C. Dickerson, Harvard Medical School and Massachusetts General Hospital; Kimoko Domoto-Reilly, University of Washington School of Medicine; David Knopman, Mayo Clinic, Rochester; Bradley F. Boeve, Mayo Clinic Rochester; Adam L. Boxer, University of California, San Francisco; John Kornak, University of California, San Francisco; Bruce L. Miller, University of California, San Francisco; William W. Seeley; University of California, San Francisco; Maria-Luisa Gorno-Tempini, University of California, San Francisco; Scott McGinnis, University of California, San Francisco; Maria Luisa Mandelli, University of California, San Francisco.

