## Supplemental material for "Distribution of White Matter Hyperintensities across Arterial Territories in Neurodegenerative Diseases"

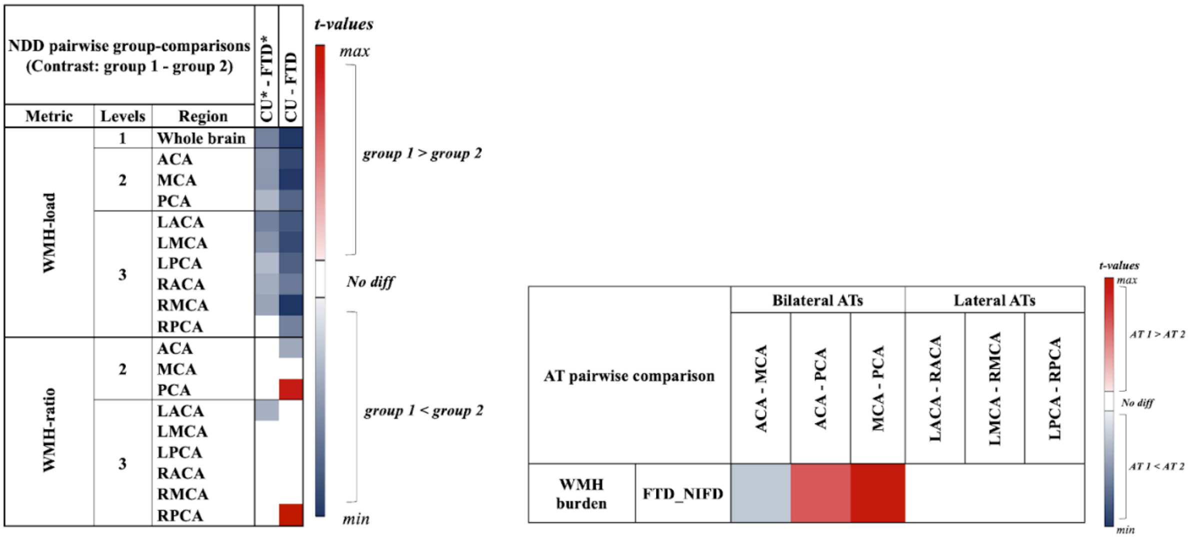


***Supplemental Fig. 1.*** *Group- and AT- comparison findings from the CU_NIFD and FTD_NIFD groups*

Group pairwise comparison results are on the left and AT pairwise comparisons on the right. To be read as Fig. X and Y, respectively.

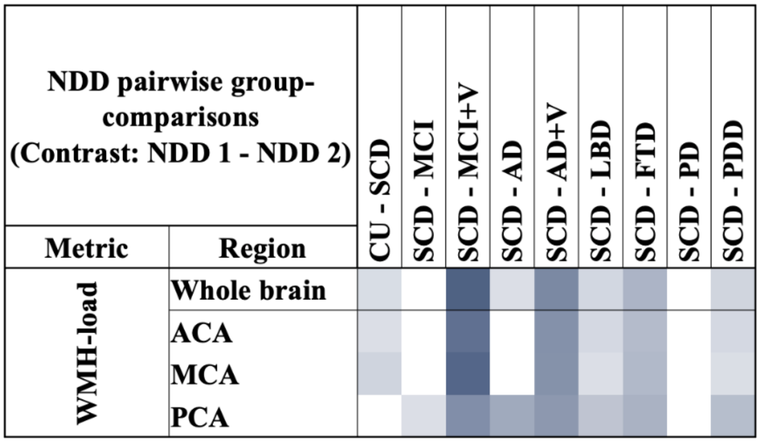


***Supplemental Fig. 2.*** AT-specific WMH burden comparisons between SCD and other clinical categories. Columns indicate pairs of contrasted groups (group 1 - group 2), each row referring to a distinct AT. The red color represents regions where group 1 has a significantly higher WMH ratio than group 2, with the gradient representing $t$-values. Similarly, the blue color represents regions where group 1 has a significantly lower WMH ratio than group 2. White cells represent contrasts that exhibit no significant statistical difference. Results are corrected for multiple comparisons using FDR.

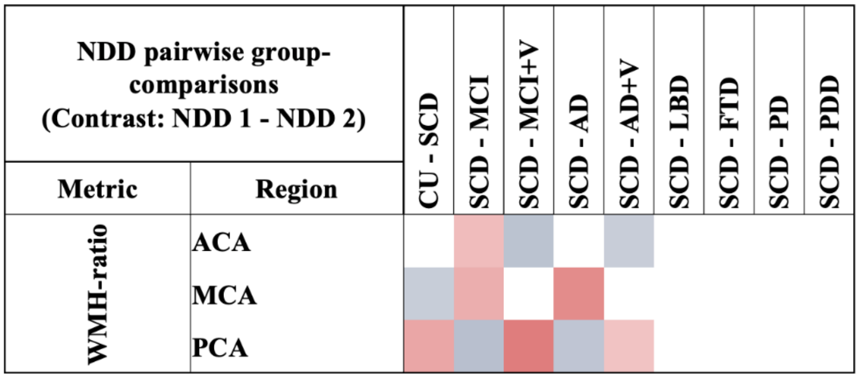


***Supplemental Fig. 3.*** *AT-specific WMH ratio comparisons between SCD and other clinical categories.* **a)** Columns indicate pairs of contrasted groups (group 1 - group 2), each row referring to a distinct AT. The red color represents regions where group 1 has a significantly *higher* WMH ratio than group 2, with the gradient representing *t*-values. Similarly, the blue color represents regions where group 1 has a significantly *lower* WMH ratio than group 2. White cells represent contrasts that exhibit no significant statistical difference. Results are corrected for multiple comparisons using FDR.

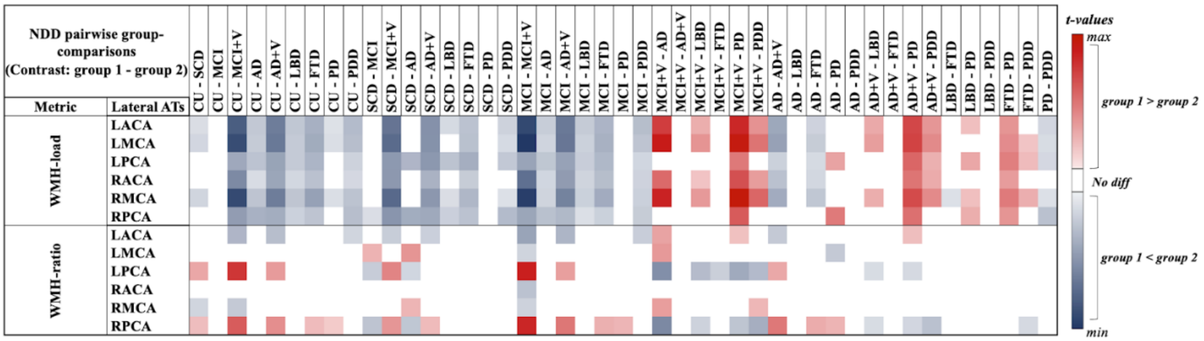


***Supplemental Fig. 4.*** *Group-comparisons in the Lateral ATs*

Columns indicate pairs of contrasted groups (group 1 - group 2), each row referring to a distinct lateral AT. The red color represents regions where group 1 has a significantly *higher* WMH-metric (burden or ratio) than group 2, with the gradient representing *t*-values. Similarly, the blue color represents regions where group 1 has a significantly *lower* WMH-metric than group 2. White cells represent contrasts that exhibit no significant statistical difference. Results are corrected for multiple comparisons using FDR.
